# Data-driven estimation of change points reveals correlation between face mask use and accelerated curtailing of the COVID-19 epidemic in Italy

**DOI:** 10.1101/2020.06.29.20141523

**Authors:** Morten Gram Pedersen, Matteo Meneghini

## Abstract

Italy was the first Western country to be seriously affected by COVID-19, and the first to implement drastic measures, which have successfully curtailed the epidemic. To understand which containment measures altered disease dynamics, we estimate change points in COVID-19 dynamics from official Italian data. We find excellent correlation between nationwide lockdown and the epidemic peak in late March 2020. Surprisingly, we find a change point in mid April, which does not correspond to national measures, but may be explained by regional interventions. Change points in regional COVID-19 dynamics correlate well with local distribution of free face masks and regional orders requiring their mandatory use. Regions with no specific interventions showed no change point during April. We speculate that widespread use of face masks and other protective means has contributed substantially to keeping the number of new Italian COVID-19 cases under control in spite of society turning towards a new normality.

## Introduction

The COVID-19 disease due to the SARS-CoV-2 coronavirus is spreading rapidly across the globe since its outbreak in China, and was declared a pandemic by the WHO on March 11, 2020. After the first severe patient was brought to the hospital of Codogno, Italy on February 20, 2020, and subsequently tested positive for COVID-19, a rapidly increasing number of patients have been identified, initially in Northern Italy and later in the rest of the country and Europe. Italy is one of the most affected European country, with ∼250.000 confirmed cases and nearly 35.000 COVID-19 related deaths, and was the first to implement drastic contain measures. The imposed restrictions, culminating with complete lockdowns, have turned out to be effective in controlling the epidemic in Italy; the number of new daily cases peaked in late March 2020 at ∼6000 and declined to ∼200 − 300 by early June. The limitations in activities were followed by milder orders and direct invitations to behavioural change, such as distribution of face masks accompanied by their mandatory use, first in the most hit regions and later nationwide. During the month of May 2020, the country reopened many activities without compromising the decay in the number of newly infected individuals. Analyzing the Italian data carefully may therefore provide important insights into the epidemiology of COVID-19, and in particular to investigate if, how and which limitations in activities and other actions affected the disease dynamics in Italy.

Effective measures slow diffusion of the disease. By identifying such change points in COVID-19 spreading [1], it is therefore possible to associate interventions that were able to modify the course of the epidemic without assuming any effect a priori. Further, such an approach may reveal wether e.g. reopening of society lead to changes in disease dynamics, or could hint at change points apparently unrelated to regulations that deserve further investigations. To find such change points, it is advantageous to use relatively simple mathematical models of infectious diseases, which can be fitted to data with a minimum number of assumptions on model parameters [1–3]. In our setting, a SIQR (susceptible – infectious – quarantined – recovered) model [4] is appropriate. In this model, identified infected individuals (cases) may be isolated, entering the “quarantined” subpopulation Q, reflecting the situation in Italy.

Preferably, one would like to have control group consisting of a matched population, but where no interventions were imposed, in order to evaluate the effects of a given intervention. Here we take of advantage of the fact among demographically similar Italian regions, some introduced specific regional containment measures at different times, whereas others did not impose any limitations beyond the nationwide rules. Due to the nationwide lockdown from March 11 to May 4, each region was basically isolated from the rest of the country during this period. We show that the model fits Italian national and regional data for the COVID-19 disease very well, and identifies change points that correlate well with national and regional containment measures, including lockdowns and widespread face mask use.

## Results

To identify change points in COVID-19 dynamics in Italy, we allow the infection rate *β* in the SIQR model to be a piecewise constant function of time, thus modelling how contain measures may affect the rate of COVID-19 transmission. The assumption of piecewise constant *β* is equivalent to the number of infectious individuals *I*(*t*) being a piecewise exponential function (under the assumption that most of the population is susceptible, *S* ≈ *N*, see Methods). Under the assumptions of the model, the daily number of new cases is then *ηI*(*t*), where *η* models the average rate with which infectious individuals become tested and quarantined, and eventually appear in the official statistics (see Methods). Hence, the log-transformed daily number of new cases is piecewise linear, allowing us to perform piecewise-linear (“segmented”) statistical modelling. This fact underlies our Data analysis and conclusions. We estimate both the time points (change points, *T*_*i*_) where the disease dynamics changes and the values of the exponential growth/decay rate *ρ* = *ρ*_*i*_ in the intervals (*T*_*i*−1_, *T*_*i*_] with *T*_0_ = 0 (Feb. 24, 2020) and *T*_max_ = 146 (July 19, 2020; last data point). Importantly, these estimates do not depend on the value of *η* (see Methods).

We obtained an good fit to the Italian data of daily COVID-19 cases (Fig. 1). The data analysis identified four change-points, separating intervals with exponential growth or decay clearly seen also in the raw data (straight lines in Fig. 1). We confront these change points to public interventions by assuming a delay from a change in the transmission rate to appearance in the official data, effectively shifting the change point backwards in time. This approach provides reasonable estimates of changes in disease dynamics [5]. We consider a 5-days delay from infection to the onset of symptoms [6, 7], followed by a variable delay from onset of symptoms to registration. This latter delay varied during the epidemic ranging 2-6 days according to the data from Italian health authorities, which grouped the data and provided the median delay for 10-days intervals [8]. Thus, the total delay from time-of-infection to observation, corresponding to the shift of change points when confronting with public interventions, is between 7 and 11 days.

**Fig 1.**
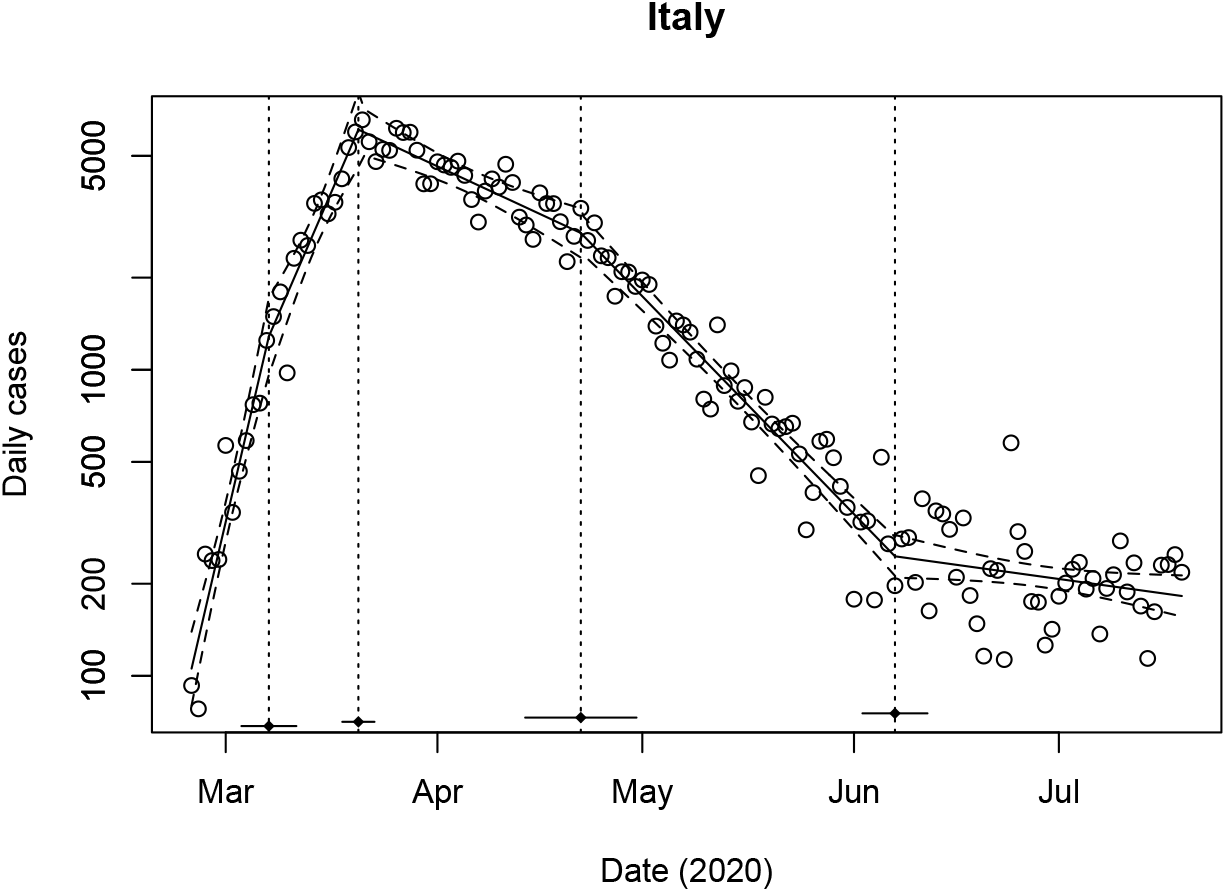
Model fit to national Italian data. The daily number of new of confirmed COVID-19 cases (circles) on logaritmic scale, and the best segmented fit (full curve) with 95 % confidence bands (dashed). The black dots and the vertical dashed lines indicate the change points *T*_1_, *T*_2_, *T*_3_ and *T*_4_, whereas the horizontal lines show the 95% confidence intervals for the change points. Estimated change points and growth rates: *T*_1_ = 12.2 days (March 7, 2020; SE 2.0 days), *T*_2_ = 25.4 days (March 20, 2020; SE 1.2 days), *T*_3_ = 58.0 days (April 21, 2020; SE 4.1 days), *T*_4_ = 104.0 days (June 6, 2020; SE 2.4 days), *ρ*_1_ = 0.221/day (SE 0.022/day), *ρ*_2_ = 0.119/day (SE 0.019/day), *ρ*_3_ = −0.024/day (SE 0.005/day), *ρ*_4_ = −0.053/day (SE 0.003/day), and *ρ*_5_ = −0.007/day (SE 0.003/day). Data from https://github.com/CSSEGISandData/COVID-19/tree/master/csse_covid_19_data/csse_covid_19_time_series.

The first change point was estimated to fall on March 7, 2020 (95% C.I. [March 3, March 11]). We note that the number of tests increased rapidly during the first weeks following the confirmation of the first Italian COVID-19 case on February 22, 2020, and the initial rapid rise might be due to testing of a pool of already infected individuals. Further, this first change point is highly dependent on a relatively few number of data points in late February – early March. Because of the related uncertainties we do not speculate further on any possible underlying cause.

The second change points was estimated to fall on March 20, 2020 (95% C.I. [March 18, March 23]), following which the growth rate became negative. For the period March 11 – 20, the median delay between onset of symptoms and reporting was 5 days [8], which should be added to the 5 days of incubation time. Thus, the change point should be compared with interventions occurring around March 10. This date is in excellent agreement with the lockdown of the Northern regions on March 8, 2020, which was followed by complete lockdown of Italy on March 11. A third change point was found at April 22, 2020 (95% C.I. [April 14, April 30]) where the decay was further accelerated. This latter change point is apparently not related to any specific nationwide containment measures. Since early June (change point June 7) the number of cases is nearly constant. We speculated that regional interventions might underlie the change in dynamics seen in April and proceeded by analyzing the data in the eight hardest hit Italian regions.

We found that all regions had a change point in late March (for Lombardy and Marche we were able to identify two, similar to what we found for the national data, but we note that the early change point depends on very few data points in late February – early March), and that Veneto, Emilia-Romagna, Piedmont, Tuscany and Liguria had another change point in mid-to-late April. In spite of reopening of society from May 4, the daily number of new cases continued to decline until early June where, in most regions, the number of new cases became so low that small fluctuations and local hotspots make the analysis less reliable. We note that there is a recent increase in the number of cases in Veneto and Emilia-Romagna, which is somewhat worrying.

Lombardy, the worst hit Italian region, showed slight reduction in the growth rate from the change point on March 3 (95% C.I. [February 29, March 7]). The number of daily cases peaked in late March corresponding to the change point identified on March 19 (95% C.I. [March 16, March 22]). Since the peak a remarkable constant exponential decline is observed. Shifting the identified change point on March 19 ten days backwards corresponds very well to date of the lockdown of Lombardy on March 8, 2020. Following the lockdown and nationwide closure of all non-essential work places on March 22, Lombardy introduced mandatory face mask use on April 4, 2020 [9]. We note that in late March – early April, the regional health care system was close to a collapse, and the data may therefore be unreliable near the peak due e.g. reduced testing capacity or longer-than-normal reporting delays, making it difficult to distinguish between the different containment measures. Reopening of society has not modified the dynamics of the epidemic in Lombardy, which – as remarked above – shows sustained exponential decay since mid-to-late March 2020.

For Veneto, the first identified change point fell on March 21 (95% C.I. [March 17, March 24]), reflecting that the number of daily new cases stabilized at ∼400 with a small decline until the second change point found at April 21 (95% C.I. [April 14, April 28]). The number of new cases then showed a marked exponential decline. As for Lombardy, the lockdown on March 8 correspond well to the first change point considering a 10 days delay. During the period April 10–19, the median delay from onset-of-symptoms to reporting was 4 days [8], corresponding to a 9 days shift of the change point. The second identified change point thus correlates well with the mandatory use of face masks from April 14 in Veneto [10].

For Tuscany we found a similar pattern with a near-constant plateau during late March – early April followed by exponential decline. The corresponding latter change point was April 16 (95% C.I. [April 8, April 24]). Shifting this date 9 days backwards in time, we find good temporal correlation with the regional order of April 6, 2020, regarding the distribution of face masks in Tuscany from April 7, anticipating mandatory use in single municipalities once the distribution had completed, and from April 20 in the entire Region [11].

Piedmont showed a slightly increasing number of daily cases in late March – early April until the change point on April 18 (95% C.I. [April 12, April 25]). This change point correlates well with the introduction of a series of strict measures in Piedmont, including the ban of gatherings of more than two persons, from April 14 [13]. Piedmont did not require face masks until May 4, 2020, but the regional government announced on April 15 that masks would become mandatory and started their distribution soon after (from April 25, [12]).

For Emilia-Romagna and Liguria the change points were, respectively, April 28 (95% C.I. [April 19, May 6]) and April 22 (95% C.I. [April 5, May 9]). Note the large uncertainty for Liguria. These change points should be shifted 9-10 days backwards in time [8], which provides good correlation with distribution of free face masks. For example, in the capital of Emilia-Romagna, Bologna, distribution of face masks started April 14 [14], and in Liguria distribution occurred from April 20 [15].

The remaining two regions Marche and Lazio correspond to the “control group” in this observational study. Neither distributed free face masks, nor required their mandatory use before the nationwide order on May 4. No other particular interventions were ordered in Marche or Lazio to contain COVID-19 spreading.

For Marche, we identified an early change point as mentioned above, and a second change point on March 19 (95% C.I. [March 17, March 22]) corresponding to the peak in the data. As for the other regions, this change point correlates well with the lockdown on March 11. A third change point was found on May 17 (95% C.I. [March 12, March 21]), followed by another on May 27 (95% C.I. [March 23, March 30]). In late May, the number of cases in Marche fell below ∼10, making the data less reliable for predicting overall dynamics. Importantly, we found no evidence of a change point during April, where the daily number of new cases showed steady exponential decay. The analysis of the data from Lazio revealed a change point on March 22 (95% C.I. [March 20, March 23]), which corresponds very well to the lockdown on March 11. A second change point was identified to fall on June 19 (95% C.I. [June 13, June 24]). As for Marche, no change point was found for the month of April.

The relevance of the above change points for fitting the data was found following model selection using ANOVA, AIC and BIC. We performed additional tests (Davies’ test [16]) against the null-hypothesis of no change in *ρ* from the peak to mid May, i.e., during March 22 – May 16. These tests confirmed that the change in slope was significant for Emilia-Romagna, Piedmont, Tuscany, Veneto (all *p* < 0.001), and Liguria (*p* = 0.015), whereas there was no evidence for a change of slope during this period for Lazio (*p* = 0.065), Lombardy (*p* = 0.88) or Marche (*p* = 1).

## Discussion and Conclusion

The importance of face mask use for controlling COVID-19 by reducing transmission from asymptomatic individuals during the reopening of societies has been widely claimed [17–21]. The Italian data provide an opportunity to investigate whether the dynamics of the epidemic had any correlation with the use of face masks, since demographically similar regions behaved differently regarding orders and distributions promoting face masks use.

To study the dynamics of COVID-19, we estimated change points [1]. We found a change point reflecting the peak in the number of new cases for all eight regions that readily explain the change point for the national data. Temporally, these change points correlate well with the lockdown imposed on March 8 – 11 depending on the region. Our analysis revealed that five of the eight analyzed regions showed change points during April that correlate temporally well with additional general containment measures (Piedmont), the introduction of mandatory face mask use (Veneto and Tuscany) and/or distribution of free face masks (Tuscany and Emilia-Romagna)

We considered alternative explanations for the acceleration of the decline of new cases seen in late April. The number of COVID-19 tests did not decline in correspondence to the identified change point, and the fraction of positive tests showed an acceleration in the decline similar to the decline seen in the number of new daily cases (Supplementary Fig. S1). Mobility data (https://www.google.com/covid19/mobility/; Supplementary Fig. S2) showed, if anything, increased activity in late compared to early April, excluding that reduced activity underlie the change. In contrast, the lockdown introduced in early March 2020 correlate with a marked drop of ∼75% in mobility in Italy compared to pre-lockdown. The weather was mild and dry in April in Italy (https://www.3bmeteo.com), with little variation in temperature during the month. Virtually no rain fell during the entire month until April 27. Thus, no abrupt change in weather was seen around mid-April, which might have been the cause of the change in dynamics. In summary, no obvious alternative explanations for the acceleration of the end of the epidemic were found.

Summarizing, this observational study showed good temporal correlation between a series of containment measures and changes in COVID-19 dynamics in Italy. Based on the excellent correspondence with estimated change points, we speculate that lockdowns were the main cause in halting the spreading of the disease, which lead to the peak in new cases seen in late March. Analyzing regional data, we were able to distinguish change points for each region that correspond well to the introduction of face mask distributions or orders in the individual regions. The reopening of society in May did not lead to any change in the decay rate (Figs. 1 and 2). Our results thus lend further support to the importance of face mask use for controlling COVID-19.

**Fig 2.**
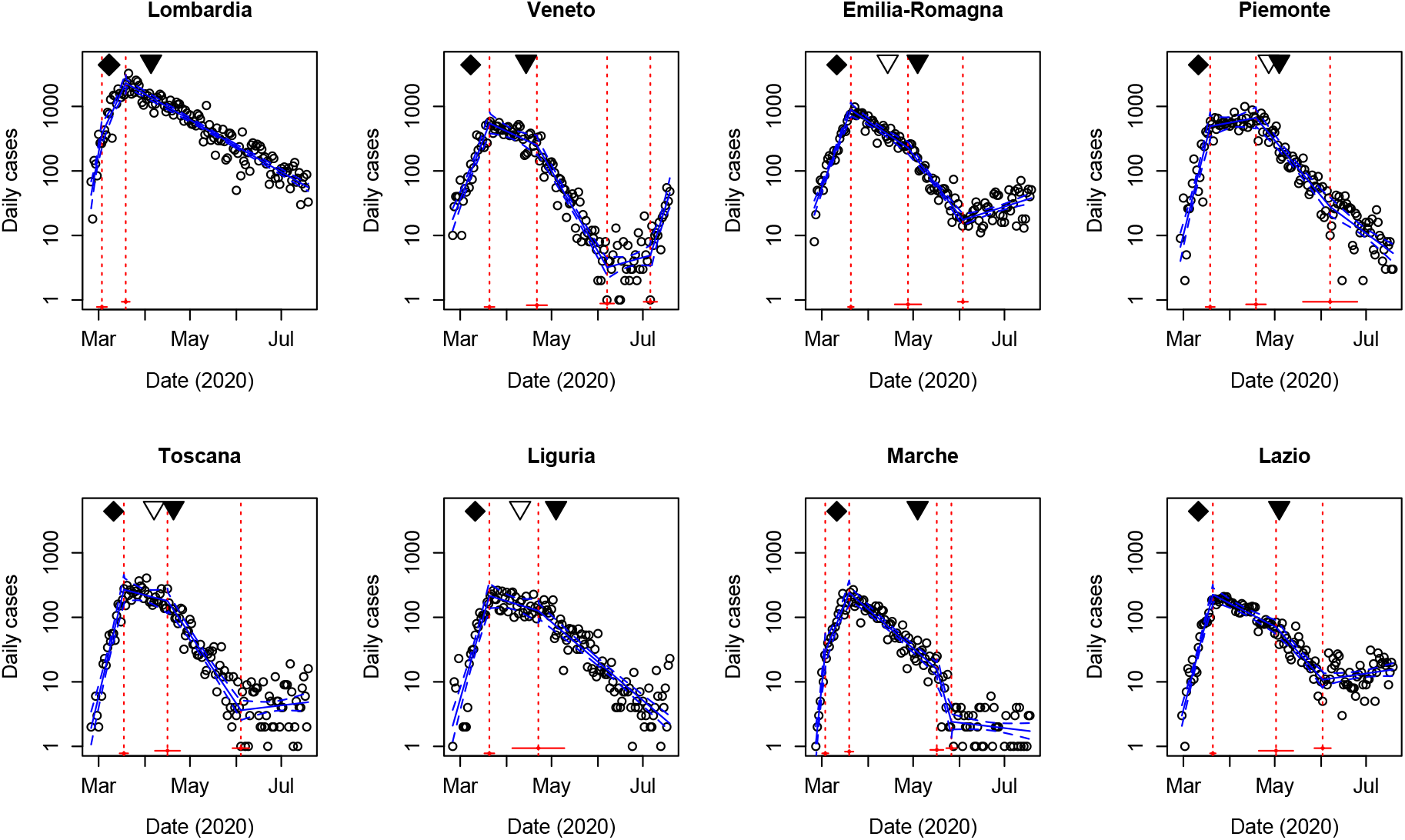
The daily number of new confirmed cases in the eight Italian regions with most COVID-19 cases, corresponding to Fig. 1B. For Emilia-Romagna, Piedmont, Tuscany and Liguria only two change points were identifiable. Data from https://github.com/CSSEGISandData/COVID-19/tree/master/csse_covid_19_data/csse_covid_19_time_series.

## Methods

We use a SIQR model [4] to describe COVID-19 dynamics in Italy, motivating our statistical analysis described below. Since we are fitting the official number of new daily cases, we unite the *Q* and *R* compartments in a “cases” compartment *C* = *Q* + *R*. The model equations are

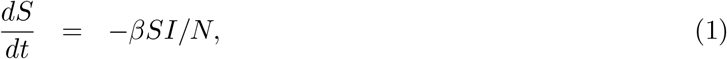

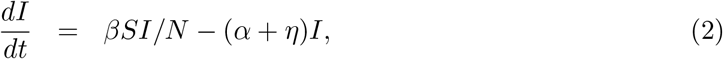

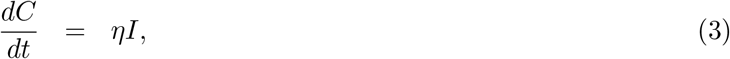

where *β* is the rate of infection, *η* models the average rate with which infectious individuals become tested and quarantined, and eventually appear in the official statistics, and *α* is the rate with which unidentified infectious individuals recover or die from the disease. We do not explicitly model the number of recovered or deceased non-identified COVID-19 patients, but only the rate *α* with which these patient become non-infectious. Further, *N* is the total number of individuals in the population, assumed constant since we are studying the early phase of the epidemic. Note that *S* + *I* + *C* ≠ *N*. To obtain such a conservation law, an additional compartment *R*_*I*_ that model recovered but non-identified patients could be added with dynamics *dR*_*I*_*/dt* = *αI*.

Since there is evidence that COVID-19 can be transmitted in the absence of symptoms [22–24], we do not include an explicit exposed-but-noninfectious (E) state, i.e., we do not consider a SEIQR model [25]. Further, from the Diamond Princess cruiseship and from the Italian village Vo’ Euganeo, it has been found that ∼50% of COVID-19-positive individuals do not develop symptoms [26, 27]. We assume that such positive but asymptomatic individuals can transmit the disease, i.e., the *I* state includes both individuals that will not develop symptoms, cases that did not develop symptoms yet, and symptomatic patient that still have not been tested positive and isolated.

In spite of the fact that ∼240,000 individuals have been found COVID-19 positive, and possibly a few millions of unidentified cases have occurred, the total number of persons that have had the infection constitute a relatively small fraction of the Italian population of ∼60 million. Thus, it is a reasonable approximation that most of the population is still susceptible, *S* ≈ *N*, and hence, as well known, the number of infected individuals follows at any time exponential growth or decay, unless parameters change.

To identify change points, we allow *β* to be a piecewise constant function of time, thus modelling how contain measures may affect the rate of COVID-19 transmission. We estimate both the time points (change points, *T*_*i*_) where *β* changes and the values of *β* = *β*_*i*_ in the intervals (*T*_*i*−1_, *T*_*i*_] with *T*_0_ = 0 (Feb. 24, 2020) and *T*_max_ = 122 (July 19, 2020; last data point). The assumption of piecewise constant *β* is equivalent to *I*(*t*) being a piecewise exponential function (under the assumption *S* ≈ *N*) with piecewise constant growth rate *ρ*_*i*_ = *β*_*i*_ − (*α* + *η*). Since *dC/dt* in proportional to *I*, also the number of new daily cases will be a piecewise exponential function, and hence the log-transformed daily increase in the number of cases, log(*dC/dt*), will be a piecewise linear function. This fact underlies our data analysis and conclusions.

We log-transform the number of newly daily cases and fit piecewise linear (“segmented”) statistical models to the data using the segmented package [28] in R [29], which provides estimates for *T*_*i*_ and *ρ*_*i*_. The number of change points was increased subsequently, and model selection was performed using ANOVA, AIC and BIC in order to obtain a parsimonious model. Residuals were verified graphically to be symmetrically distributed around zero, homoscedastic, and show no sign of autocorrelation. Davies’ test for “no change in slope” was applied as indicated in the test for confirming the model selection results regarding change points in April – early May. *p*-values below 0.05 were considered significant.

## Data availablity

The data that support the findings of this study are publicly available at https://github.com/CSSEGISandData/COVID-19/tree/master/csse_covid_19_data/csse_covid_19_time_series (Figs. 1, 2, S1) and at https://www.google.com/covid19/mobility/ (Fig. S2).

## Code availablity

Computer code used for the analysis of the data is available from the corresponding author upon reasonable request.

## Authors contributions

MGP and MM conceived research and discussed all results. MGP developed the theoretical framework, analyzed data, performed parameter estimation, and wrote the paper. MM commented drafts and approved the final version of the paper.

## References

1. Dehning J, Zierenberg J, Spitzner FP, Wibral M, Neto JP, Wilczek M, et al. Inferring change points in the spread of COVID-19 reveals the effectiveness of interventions. Science. 2020;doi:10.1126/science.abb9789.

2. Hethcote HW. The mathematics of infectious diseases. SIAM review. 2000;42(4):599–653.

3. Eubank S, Eckstrand I, Lewis B, Venkatramanan S, Marathe M, Barrett CL. Commentary on Ferguson, et al., “Impact of Non-pharmaceutical Interventions (NPIs) to Reduce COVID-19 Mortality and Healthcare Demand”. Bull Math Biol. 2020;82(4):52. doi:10.1007/s11538-020-00726-x.

4. Hethcote H, Zhien M, Shengbing L. Effects of quarantine in six endemic models for infectious diseases. Mathematical biosciences. 2002;180(1-2):141–160.

5. Gostic KM, McGough L, Baskerville E, Abbott S, Joshi K, Tedijanto C, et al. Practical considerations for measuring the effective reproductive number, Rt. medRxiv. 2020;.

6. Li Q, Guan X, Wu P, Wang X, Zhou L, Tong Y, et al. Early Transmission Dynamics in Wuhan, China, of Novel Coronavirus-Infected Pneumonia. N Engl J Med. 2020;382(13):1199–1207. doi:10.1056/NEJMoa2001316.

7. Lauer SA, Grantz KH, Bi Q, Jones FK, Zheng Q, Meredith HR, et al. The Incubation Period of Coronavirus Disease 2019 (COVID-19) From Publicly Reported Confirmed Cases: Estimation and Application. Ann Intern Med. 2020;172(9):577–582. doi:10.7326/M20-0504.

8. Istituto Superiore di Sanità (ISS), Rome. Epidemia COVID-19 Aggiornamento nazionale 23 giugno 2020; 2020. Available from: https://www.epicentro.iss.it/coronavirus/bollettino/Bollettino-sorveglianza-integrata-COVID-19_23-giugno-2020.pdf [cited July 1, 2020].

9. https://www.ansa.it/documents/1586025090469_Ordinanza_Lombardia_203004044_obbligo_mascherine.pdf;.

10. https://bur.regione.veneto.it/BurvServices/pubblica/DettaglioOrdinanzaPGR.aspx?id=418285;.

11. https://www.comunesgv.it/content/uploads/2020/04/Ordinanza_del_Presidente_n.26_del_06-04-2020.pdf;.

12. https://www.regione.piemonte.it/web/pinforma/notizie/5-milioni-mascherine-per-tutti-piemontesi;.

13. https://www.regione.piemonte.it/web/sites/default/files/media/documenti/2020-04/decreto_presidente_della_giunta_regionale_n._43_-_13_aprile_2020.pdf;.

14. http://www.comune.bologna.it/news/coronavirus-mascherine;.

15. https://www.lastampa.it/savona/2020/04/19/news/le-poste-riprendono-la-distribuzione-d38738004;.

16. Davies RB. Hypothesis testing when a nuisance parameter is present only under the alternative: linear model case. Biometrika. 2002; p. 484–489.

17. Prather KA, Wang CC, Schooley RT. Reducing transmission of SARS-CoV-2. Science. 2020;368(6498):1422–1424. doi:10.1126/science.abc6197.

18. Li R, Pei S, Chen B, Song Y, Zhang T, Yang W, et al. Substantial undocumented infection facilitates the rapid dissemination of novel coronavirus (SARS-CoV-2). Science. 2020;368(6490):489–493. doi:10.1126/science.abb3221.

19. Chu DK, Akl EA, Duda S, Solo K, Yaacoub S, Schünemann HJ, et al. Physical distancing, face masks, and eye protection to prevent person-to-person transmission of SARS-CoV-2 and COVID-19: a systematic review and meta-analysis. Lancet. 2020;395(10242):1973–1987. doi:10.1016/S0140-6736(20)31142-9.

20. Leung NHL, Chu DKW, Shiu EYC, Chan KH, McDevitt JJ, Hau BJP, et al. Respiratory virus shedding in exhaled breath and efficacy of face masks. Nat Med. 2020;26(5):676–680. doi:10.1038/s41591-020-0843-2.

21. Eikenberry SE, Mancuso M, Iboi E, Phan T, Eikenberry K, Kuang Y, et al. To mask or not to mask: Modeling the potential for face mask use by the general public to curtail the COVID-19 pandemic. Infectious Disease Modelling. 2020;5:293–308. doi:10.1016/j.idm.2020.04.001.

22. Bai Y, Yao L, Wei T, Tian F, Jin DY, Chen L, et al. Presumed Asymptomatic Carrier Transmission of COVID-19. JAMA. 2020;323:1406–1407. doi:10.1001/jama.2020.2565.

23. Rothe C, Schunk M, Sothmann P, Bretzel G, Froeschl G, Wallrauch C, et al. Transmission of 2019-nCoV infection from an asymptomatic contact in Germany. New England Journal of Medicine. 2020;382:970–971.

24. He X, Lau EH, Wu P, Deng X, Wang J, Hao X, et al. Temporal dynamics in viral shedding and transmissibility of COVID-19. Nature medicine. 2020;26(5):672–675.

25. Jumpen W, Wiwatanapataphee B, Wu Y, Tang I. A SEIQR model for pandemic influenza and its parameter identification. International Journal of Pure and Applied Mathematics. 2009;52(2):247–265.

26. National Institute of Infectious Diseases Japan. Field Briefing: Diamond Princess COVID-19 Cases, 20 Feb Update;. Available from: https://www.niid.go.jp/niid/en/2019-ncov-e/9417-covid-dp-fe-02.html.

27. Lavezzo E, Franchin E, Ciavarella C, Cuomo-Dannenburg G, Barzon L, Del Vecchio C, et al. Suppression of a SARS-CoV-2 outbreak in the Italian municipality of Vo’. Nature. 2020;doi:10.1038/s41586-020-2488-1.

28. Muggeo VM. Estimating regression models with unknown break-points. Statistics in Medicine. 2003;22(19):3055–3071.

29. R Core Team. R: A Language and Environment for Statistical Computing; 2016. Available from: https://www.R-project.org/.

